# Pre-treatment clinical behavioral and blood leukocyte gene expression patterns predict rate of change in response to early intervention in autism

**DOI:** 10.1101/2020.12.21.20248674

**Authors:** Michael V. Lombardo, Elena Maria Busuoli, Laura Schreibman, Aubyn C. Stahmer, Tiziano Pramparo, Isotta Landi, Veronica Mandelli, Natasha Bertelsen, Cynthia Carter Barnes, Vahid Gazestani, Linda Lopez, Elizabeth C. Bacon, Eric Courchesne, Karen Pierce

## Abstract

Early detection and intervention are believed to be key to facilitating better outcomes in children with autism, yet the impact of age at treatment start on outcome is poorly understood. While clinical traits such as language ability have been shown to predict treatment outcome, whether or not and how information at the genomic level can predict treatment outcome is unknown. Leveraging a cohort of toddlers with autism who all received the same standardized intervention at a very young age and provided a blood sample, here we find that very early treatment engagement (i.e., < 24 months) leads to greater gains while controlling for time in treatment. Pre-treatment clinical behavioral measures predicts 21% of the variance in the rate of skill growth during early intervention. Pre-treatment blood leukocyte gene expression patterns also predicts rate of skill growth, accounting for 13% of the variance treatment slopes. Results indicated that 295 genes can be prioritized as driving this effect. These treatment-relevant genes highly interact at the protein level, are enriched for differentially histone acetylated genes in autism post-mortem cortical tissue, and are normatively highly expressed in variety of subcortical and cortical areas important for social-communication and language development. This work indicates for the first time that gene expression can predict the rate of early intervention response and that a key biological factor linked to treatment outcome could be the susceptibility for epigenetic change via mechanisms such as histone acetylation.

---

Early detection and intervention in autism spectrum disorder (ASD) are topics of paramount importance because of the enormous potential to capitalize on the brain’s enhanced plasticity during early development as a mechanism to positively impact outcomes (*1*). While it is becoming increasingly clear that the biology of autism starts in early prenatal development (*2, 3*) and that early behavioral signs begin to manifest before 18 months (*4*–*6*), the mean age of diagnosis is still lagging far behind at 3-4 years of age (*7, 8*). In contrast to this reality, we have recently shown that diagnostic stability at much earlier ages is indeed high (*9*) and thus, the ability to detect and start treatment earlier is feasible. Some have suggested that detection and intervention before 24 months is key in order to capitalize on early neuroplasticity to facilitate optimal outcomes (*10, 11*). The impact of starting intervention earlier would likely be more total positive gains for the child (indexed by absolute level of improvement). However, a less obvious, but perhaps equally important effect of earlier intervention, could be a decrease in the variability of treatment responses at a group-level. If this were the case, the reduction in treatment response variability might allow for more precise predictions about treatment outcomes.

Understanding the ingredients that moderate and predict early intervention treatment response is of the utmost importance, especially given the current state of the field, where there is notably large heterogeneity in how individuals may respond to a treatment (*12*). While the field has noted that some early interventions have impact at a group level (*13, 14*), what is less clear is how to predict an individual’s specific response to the treatment and how to make that prediction before treatment begins. Understanding individual-level predictors of treatment response, particularly pre-treatment individual characteristics, is a key objective for precision medicine (*15, 16*) applied to autism. Ideally, we would like to know what child-specific characteristics are present before an intervention starts, in order to help us optimally predict how that specific intervention may affect the child. There are indications that some pre-treatment characteristics such as level of play, language, social cognitive abilities, IQ, ASD symptom severity, and adaptive behavior may be important for moderating treatment response (*10, 17*–*22*). In contrast to the many clinical studies that have been carried out on these phenotypic characteristics, biological moderators of treatment responses remain largely unknown, leaving open the possibility that individual intrinsic biological characteristics of a child may also moderate their response to treatment. If we better understood such treatment-relevant and individualized biological characteristics, this might yield unique insights into how and why some treatments work better for some individuals, but not others.

In this work, we examine the effect of relatively early (<24 months) versus later (≥24 months) treatment start and how this may affect total gains and variability in treatment response. We also investigate whether pre-treatment standardized clinical behavioral measures and blood leukocyte gene expression patterns moderate how quickly an individual will respond to early intervention. We operationalize treatment response here as the rate at which children respond over time and will refer to this concept from here on as ‘treatment slopes’. Blood leukocyte gene expression offers up a powerful in-vivo alternative to clinical behavioral measures, as it helps to map out biological mechanisms of brain-relevance but in a peripheral non-neural tissue. While the brain is largely an inaccessible tissue to assay mechanisms like gene expression in living patients, blood leukocyte gene expression has revealed a number of interesting brain-relevant characteristics that can be related to different phenotypes in living patients. Leukocyte expression patterns can be used in a classifier to predict diagnostic status (*23*), correlate with total brain size (*24*), and are related to large-scale functional neural systems response to speech (*25*), the patterning of thickness and surface area in the cerebral cortex (*26*), and social symptom severity (*27*). Differentially expressed genes in blood leukocytes are part of extended gene networks that are linked to highly penetrant ASD-related mutations (*27*). Another revelation is that blood leukocyte genes associated with autism tend to be within a class of broadly expressed genes that are highly expressed in the brain and many other tissues (*25*). Broadly expressed genes are one class of important ASD-associated genes that primarily have peak levels of expression during prenatal development (*3, 28*). Given the sensitivity of blood leukocyte gene expression activity as a tool for assessing the living biology behind ASD toddlers (*2*), we reasoned that there may be pre-treatment gene expression patterns in ASD toddlers that may be predictive of treatment slopes.

In this study, we used least absolute shrinkage and selection operator (LASSO) regression (*29*) to model how clinical behavioral measures or gene expression patterns may be predictive of treatment slopes. LASSO is an important modeling strategy here for its use of L1 regularization, which acts to penalize largely uninformative features and results in a sparse solution that allows the user to isolate the specific subset of features that are highly predictive. To better understand treatment-relevant genes, we ran further analyses to test if these genes highly interact at the protein level, whether they overlap with known ASD-related genomic and epigenomic mechanisms, and how they are expressed spatially throughout the brain.

## Methods

### Participants

This study was approved by the Institutional Review Board at the University of California, San Diego. Participants and families in this study were recruited as part of a larger multidisciplinary research project examining early neurobiological features and development of ASD at the University of California, San Diego. Toddlers with high likelihood for an ASD diagnosis were identified from one of two sources: general community referral (e.g., website or outside agency) and a population-based screening method called Get SET Early (*6, 30*). Using this population-based screening approach, toddlers with high likelihood for an ASD diagnosis as young as 12 months were identified in pediatric offices with a broadband screening instrument – the Communication and Symbolic Behavior Scales-Developmental Profile Infant Toddler Checklist (*31*). Toddlers were evaluated and tracked every six months until their third birthday when a final diagnosis was given. Licensed clinicians with expertise evaluating and diagnosing ASD in toddlers made final diagnoses based on clinical judgment and by incorporating criteria for ASD on the Autism Diagnostic Observation Schedule (ADOS) (*9, 32*). Toddlers who were determined to be high likelihood for ASD were offered intervention through our UCSD treatment program. Seventy-two families were referred for intervention, and 49 families chose to receive treatment in our program. Of the 49 children who received treatment from our program, 41 children (33 male, 8 female, mean age at start of treatment = 22.77 months, SD age = 4.08, range = 13–27 months) also had a blood sample taken before the start of treatment and were therefore included in analyses for this work. Additional participant pre-treatment clinical information can be found in Table 1. Data from this study have been previously reported in Bacon et al., (*33*) although this prior paper only focused on treatment and clinical behavioral data and did not examine gene expression.

**Table 1:** Pre-treatment clinical characteristics. Abbreviations: ADOS, autism diagnostic interview schedule; MSEL, Mullen Scales of Early Learning; VABS, Vineland Adaptive Behavior Scales; SC, social-communication; RRB, restrictive repetitive behaviors; EL, expressive language; RL, receptive language; VR, visual reception; FM, fine motor; ELC, early learning composite; SD, standard deviation.

|  | **Male** | **Female** |
| --- | --- | --- |
| **Sex** | 33 | 8 |
|  | **Mean** | **SD** |
| **Age at Treatment Intake** | 22.78 | 4.03 |
| **ADOS Total** | 14.27 | 6.02 |
| **ADOS SC** | 12.17 | 5.41 |
| **ADOS RRB** | 6.17 | 2.11 |
| **MSEL EL** | 31.39 | 10.62 |
| **MSEL RL** | 29.10 | 11.22 |
| **MSEL VR** | 41.29 | 9.65 |
| **MSEL FM** | 41.63 | 11.38 |
| **MSEL ELC** | 73.83 | 15.31 |
| **VABS Communication** | 77.12 | 14.38 |
| **VABS Socialization** | 85.51 | 12.52 |
| **VABS Daily Living Skills** | 89.05 | 11.99 |
| **VABS Motor** | 94.83 | 11.98 |
| **VABS Adaptive Behavior Composite** | 84.27 | 12.34 |

### Early intervention program

In order to reduce confounds that could be associated with differences associated with treatment type and administration, all toddlers received the same in-home treatment programming using the Strategies for Teaching Based on Autism Research (STAR) curriculum (*34*). The STAR program is a comprehensive behavioral intervention program with a curriculum designed specifically for children with ASD and includes instructional strategies of Discrete Trial Training (*35*–*37*), Pivotal Response Training (*38, 39*), and teaching in Functional Routines (*40, 41*). In an effort to improve the developmental appropriateness of the curriculum for these very young children, the STAR curriculum was augmented with developmental approaches applied through Project ImPACT. Project ImPACT is a manualized curriculum developed by Ingersoll and Dvortcsak (*42*) used to target social-communication goals in young children with ASD. Project ImPACT focuses on the relationship between adult responsivity and children’s social-communicative development. In the Project ImPACT curriculum, an early childhood interventionist (ECI) combines naturalistic behavioral strategies and developmental strategies. For example, the interventionist would respond to all communicative attempts by the child as if they were purposeful and recast expanded communication to facilitate communicative growth.

### Treatment delivery

Each child received approximately 6–12 hours per week of direct one-on-one intervention with a trained ECI at home until 36 months of age. ECIs were bachelor’s degree or undergraduate-level research assistants with previous experience with young children with ASD. Each ECI received extensive didactic and hands-on training in behavioral principles and the STAR and TSC programs discussed above. Fidelity of implementation was reached for each intervention strategy as determined by using all components of the intervention correctly at least 80% of the time across two different children and monitored for maintenance. Programs were developed and supervised by master’s degree-level clinicians (i.e., in-home coordinator) experienced in ASD, with oversight from two doctorate-level clinical psychologists with extensive experience in early behavioral intervention for this population. In addition, parent coaching was provided throughout the course of participation.

### Treatment outcome measure

The Adapted Student Learning Profile (aSLP) is a curriculum-based assessment for determining student learning goals and was adapted from the STAR curriculum to include additional goals from the TSC curriculum (see (*34, 42*)). The aSLP provides an extensive list of skills targeted in the STAR and TSC curricula and allows for the assessor to indicate the child’s performance level on each skill across six domains, receptive language, expressive language, spontaneous language, functional routines, pre-academic concepts, and play and social interaction concepts. Data were analyzed using total aSLP score across all domains rather than separate domain aSLP scores. The aSLP is administered by presenting each item up to five times to the child and observing the child’s response. This is conducted in a structured format, and no teaching was done during the assessment. The assessor then rates the child’s response, indicating if the child did not demonstrate the skill or showed partial demonstration of the skill or mastery of the skill. The entire aSLP takes about 30–45 minutes to complete. Each child’s in-home coordinator completed an aSLP at intake and every three months thereafter to determine performance and progress. A child’s performance on the treatment was estimated by the subject-specific slope estimated in a linear mixed effect model for modeling on the longitudinal aSLP scores (see section on developmental trajectory analyses).

### Pre-treatment clinical behavioral measures

Pre-treatment clinical behavioral measures were collected to characterize the sample and utilized for analyzing how predictive such pre-treatment clinical measures (measured at treatment start) were of treatment slopes. The clinical measures analyzed were the Mullen Scales of Early Learning (MSEL), the Vineland Adaptive Behavior Scales (2nd edition; VABS) and the ADOS. The MSEL assesses developmental functioning of children between birth and 68 months (*43*). An examiner measures child functioning level through a series of play-like tasks over five domains, gross motor, fine motor, receptive language, expressive language, and visual reception skills. For each scale, the assessment derives a T-score with a mean of 50 and standard deviation of 10, a percentile score, and an age equivalent score indicating at what developmental age the child is performing. An early learning composite (ELC) score is calculated from the total of scores on all scales (with the exception of the gross motor scale) with a mean of 100 and standard deviation of 15. The VABS provides a measure of adaptive skills used to cope with challenges of daily living (*44*). A caregiver completes a questionnaire regarding the individual’s current level of functioning across five domains: communication, daily living skills, socialization, motor skills, and maladaptive behavior. All scales use standard scores with a mean of 100 and a standard deviation of 15, a percentile score, and an age equivalent score indicating at what developmental age the individual is performing. Scores on all scales are combined to obtain an overall adaptive behavior composite (ABC) with a mean of 100 and a standard deviation of 15.

### Developmental trajectory analyses

To estimate aSLP trajectories for each toddler we used a linear mixed effect model to estimate longitudinal subject-specific intercepts and slopes as random effects. The subject-specific slopes (from here on called ‘treatment slopes’) estimated from this model were extracted and used as the primary treatment outcome measure to be predicted by pre-treatment gene expression or clinical measures. These analyses were computed using the *lme* function from the *nlme* library in R.

To better understand the effects of age at treatment start, we used 24 months as the cutoff point for distinguishing very early versus later treatment start. This very early versus later distinction at 24 months was made given that it is considered that the first 24 months of life are the critical early window for when early intervention could have most impact (*10, 11*). Linear mixed effect models were used to examine differences on the aSLP as a function of very early (<24 months) vs later (≥24 months) start group. We also investigated how variability in treatment slopes may differ between very early versus later start groups by computing the standard deviation of treatment slopes within each group and then quantifying the difference in standard deviation, computed as the difference score between later versus very early start groups. To test the standard deviation difference between groups against the null hypothesis of no difference in standard deviation difference score, we computed standard deviation difference scores over 10,000 random permutations of the very early or later start group labels, to derive a null distribution of standard deviation difference scores. A p-value was then computed as the percentage of times under the null distribution that a standard deviation difference score was greater than or equal to the actual standard deviation difference score.

### Blood sample collection, RNA extraction, quality control and samples preparation

Four to six milliliters of blood was collected into EDTA-coated tubes from toddlers on visits when they had no fever, cold, flu, infections or other illnesses, or use of medications for illnesses 72 hours prior blood draw. Temperature was also taken at the time of blood draw. Blood samples were passed over a LeukoLOCK filter (Ambion, Austin, TX, USA) to capture and stabilize leukocytes and immediately placed in a -20°C freezer. Total RNA was extracted following standard procedures and manufacturer’s instructions (Ambion, Austin, TX, USA). LeukoLOCK disks (Ambion Cat #1933) were freed from RNA-later and Tri-reagent (Ambion Cat #9738) was used to flush out the captured lymphocyte and lyse the cells. RNA was subsequently precipitated with ethanol and purified through washing and cartridge-based steps. The quality of mRNA samples was quantified by the RNA Integrity Number (RIN), values of 7.0 or greater were considered acceptable (*45*), and all processed RNA samples passed RIN quality control. Quantification of RNA was performed using Nanodrop (Thermo Scientific, Wilmington, DE, USA). Samples were prepped in 96-well plates at the concentration of 25 ng/µl.

### Gene expression and data processing

RNA was assayed at Scripps Genomic Medicine (La Jolla, CA, USA) for labeling, hybridization, and scanning using the Illumina BeadChips pipeline (Illumina, San Diego, CA, USA) per the manufacturer’s instruction. All arrays were scanned with the Illumina BeadArray Reader and read into Illumina GenomeStudio software (version 1.1.1). Raw data was exported from Illumina GenomeStudio, and data pre-processing was performed using the lumi package (*46*) for R (http://www.R-project.org) and Bioconductor (https://www.bioconductor.org) (*47*). Raw and normalized data are part of larger sets deposited in the Gene Expression Omnibus database (GSE42133; GSE111175).

### Patient gene expression dataset

A larger primary dataset of blood leukocyte gene expression was available from 383 samples from 314 toddlers within the UC San Diego cohort, with the age range of 1-to-4 years old. The samples were assayed using the Illumina microarray platform on three batches. The datasets were combined by matching the Illumina Probe ID and probe nucleotide sequences. The final set included a total of 20,194 gene probes. Quality control analysis was performed to identify and remove 23 outlier samples from the dataset. Samples were marked as outlier if they showed low signal intensity (average signal two standard deviations lower than the overall mean), deviant pairwise correlations, deviant cumulative distributions, deviant multi-dimensional scaling plots, or poor hierarchical clustering, as described elsewhere (*24*). The high-quality dataset included 360 samples from 299 toddlers. High reproducibility was observed across technical replicates (mean Spearman correlation of 0.97 and median of 0.98). Thus, we randomly removed one of each of two technical replicates from the primary dataset. From the subjects in the larger primary dataset, a total of n=41 also had treatment data; n=36 from the Illumina HT12 platform along with n=5 from the Illumina WG6 platform were used in this study. The 20,194 probes were quantile normalized and then variance filtered to leave the top 50% of highly varying probes (i.e. 10,097 probes). Treatment slopes were slightly different as a function of batch (*F*(*2,35*) = 3.44, *p* = 0.04), but were not different at age at blood sampling (*F*(*1,35*) = 0.001, *p* = 0.97), sex (*F*(*1,35*) = 2.09, *p* = 0.15) or RIN (*F*(*1,35*) = 0.22, *p* = 0.63). Removal of variance associated with batch, sex and RIN was achieved by using linear model to estimate these effects in the training set of each cross validation fold. This model computed on the training set was then applied to the test set for removing variance such covariates.

### Predictive modeling of treatment slopes

To predict individual differences in treatment slopes we used a LASSO regression model (*29*) which used as predictors either multivariate pre-treatment gene expression or clinical measures. LASSO uses L1 regularization (controlled by the lamba (λ) parameter) to shrink beta coefficients of uninformative features and thus reduce or effectively remove the influence of such features on the model. This feature is important for our purposes as we seek to compute a model that predicts treatment slopes but also informs us as to which features (e.g., genes or clinical measures) are most important for the model. For all LASSO modeling to assess the model’s predictive utility we used leave-one-out cross validation (CV) to partition the data into training and test sets. Within the training set, a 10-fold CV loop is used to estimate the optimal lambda parameter for the model. Cross validated mean squared error (MSE) and R^2^ were computed to evaluate the predictive value of the model. We also used permutation tests (1000 permutations) to randomly shuffle treatment slopes and construct a null distribution of MSE values under the null hypothesis. This null MSE distribution was used to compute a p-value, defined as the proportion of times under the null distribution where an MSE value was as low or lower than the observed MSE value with unpermuted treatment slopes.

### Protein-protein interaction analysis

The resulting gene list from the LASSO model predicting treatment slopes was then tested for evidence of protein-protein interactions (PPI). This analysis was achieved using the STRING database (https://string-db.org), with all parameters set to the STRING defaults (using all interaction sources and confidence interaction scores of 0.4 or higher). STRING also outputs enrichment results for Gene Ontology, Reactome, KEGG, and UniProt databases.

### Autism-associated gene set enrichment analyses

To better link the set of treatment-relevant genes prioritized by the LASSO model, we tested this gene set for enrichment with other lists of genes known from the literature to be associated with autism. For autism-associated genetic mutations we used genes from SFARI Gene (https://gene.sfari.org) (*48*) in categories S, 1, 2, and 3 (October 2020 release). For genes with evidence of dysregulated expression in post-mortem cortical tissue we used differentially expressed gene lists from Gandal et al., (*49*). At the epigenetic level, we also analyzed genes with evidence for differential histone acetylation in autism in post-mortem prefrontal and temporal cortex tissue (*50*).

### Spatial gene expression analyses

To get a better idea of the brain regions that are likely to be maximally affected by treatment-relevant genes prioritized by the LASSO model, we examined how these genes were spatially expressed across the brain using the Allen Institute Human Brain atlas (*51*). Whole-brain gene expression maps for treatment-relevant genes were downloaded in MNI space from https://neurosynth.org/genes/. These gene maps were then input into a whole-brain one-sample t-test computed in SPM12 (https://www.fil.ion.ucl.ac.uk/spm/software/spm12/). Thresholding for multiple comparisons was achieved with voxel-wise FDR correction set to q<0.05.

### Data and code availability

Analysis code is available at https://github.com/IIT-LAND/gex_treatment_slopes. Data are available from the National Institute of Mental Health Data Archive (NDA) (https://nda.nih.gov). Raw and normalized blood gene expression data are also deposited in Gene Expression Omnibus (GEO) (GSE42133; GSE111175).

## Results

### Differences between early versus late treatment start groups

A total of n=41 toddlers were considered in all further analyses given that they had both gene expression and treatment data. In a first analysis we examined whether individuals with a very early start to treatment (i.e. <24 months) would result in better outcomes than those who started treatment later (i.e. >= 24 months). This early versus late distinction at 24 months was made given that it is considered that the first 24 months of life are the critical early window for when early intervention could have most impact (*10, 11*). For this analysis, we used a linear mixed effect model that modeled treatment start group (Very Early, <24 months of age at the start of treatment; Later, >=24 months of age at the start of treatment), age, the interaction between age and treatment start group, and number of days in treatment as fixed effects and subject-specific slopes and intercepts as random effects. Main effects were observed for age (*F* =130.34, *p* = 2.22e-16) and treatment start group (*F* = 25.39, *p* = 1.17e-5), but there was no interaction between age and treatment start group (*F* = 1.23, *p* = 0.26) (Fig. 1A). These results indicate that individuals starting treatment before 24 months of age have larger total treatment gains than those who start treatment relatively later (after 24 months) and that these effects cannot be explained by factors such as the duration of time in treatment. However, the lack of an age-by-group interaction in predicting treatment slopes indicates that there are no differences in the steepness of the trajectories between early versus late start groups.

**Fig. 1:**
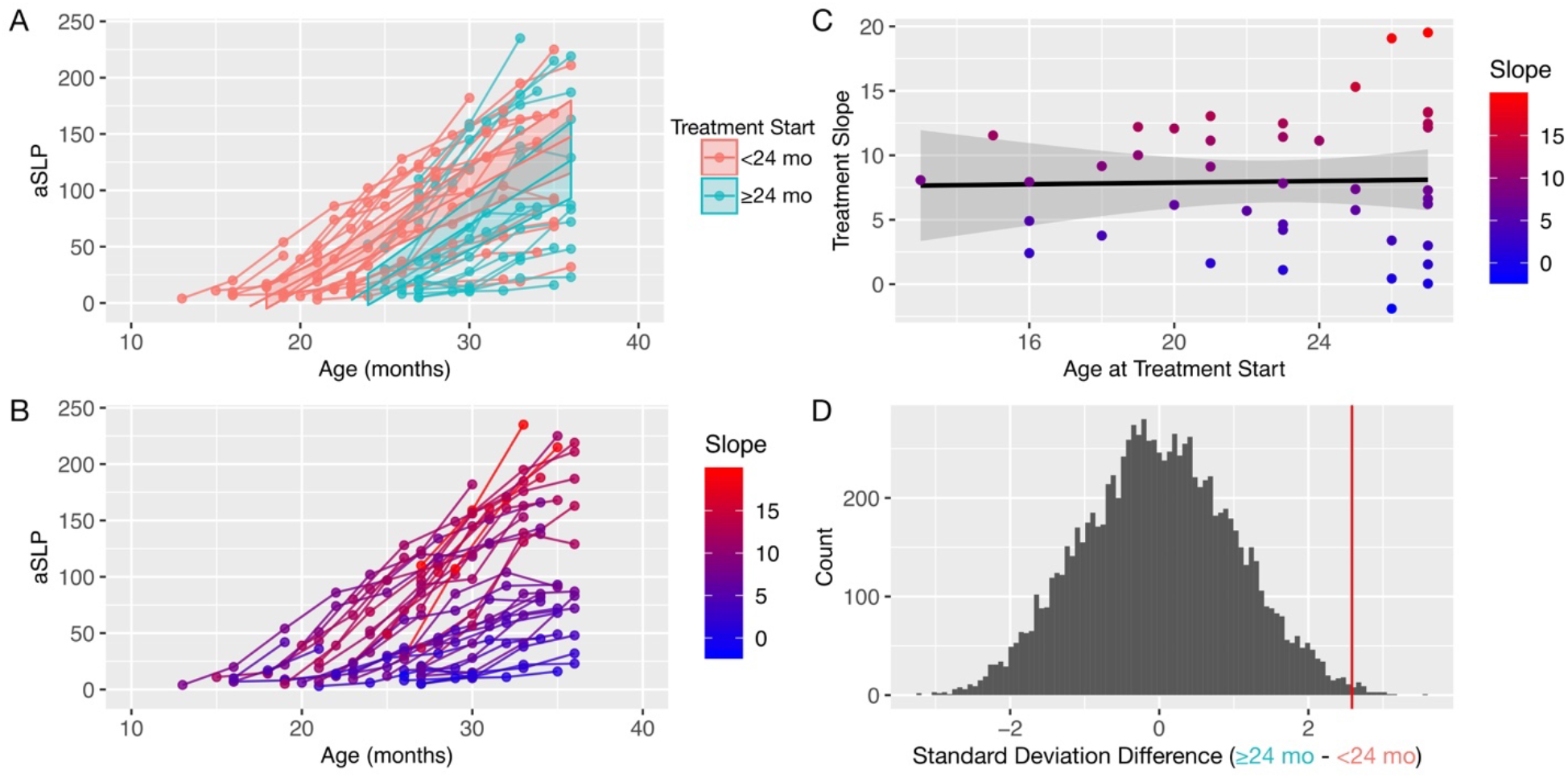
Treatment slopes and relationship with age at treatment start. Panels A and B show trajectories of skill acquisition on the Adapted Student Learning Profile (aSLP) treatment outcome measure. Panel A shows these trajectories for treatment start groups defined by age at treatment start as either very early (<24 months, pink) or relatively later (≥24 months, turquoise). Panel B shows the trajectories but with each individual’s data now colored by treatment slopes (colored from blue to red) estimated from a linear mixed effect model. Higher slopes indicate steeper trajectories and thus faster rate of skill growth over time whereas relatively lower slopes indicate less steep trajectories that can be interpreted as relatively slower rates of skill growth over time. Panel C shows treatment slopes for each individual as a function of age at treatment start (color indicates treatment slopes, as shown in panel B). Variability in treatment slopes becomes markedly larger when age of treatment start occurs after 24 months of age. Panel D shows a null distribution of difference in standard deviations over 10,000 permutations of random labelings of later (≥24 months) vs very early (<24 months) groups. The actual difference in standard deviation between later vs very early start groups is shown by the vertical red line.

While the steepness of treatment slopes do not heavily differ on-average between early versus late start groups, it is noteworthy that where the two groups do differ is on the variability in treatment slopes. Figure 1B-C shows a clear distinction between the late start group showing markedly more variable treatment slopes than the early start group. A permutation test further verified that the actual difference in standard deviations between late versus early start groups is highly significant relative to what this standard deviation difference would be under random group labeling (p = 0.004) (Fig. 1D). This result indicates that while treatment slopes remain relatively consistent in their variability before 24 months, after 24 months treatment slopes become much more variable.

### Prediction of treatment slopes with pre-treatment clinical measures

We next examined if pre-treatment clinical behavioral measures could be predictive of treatment slopes. A LASSO model that included all pre-treatment ADOS, Mullen, and VABS subscales was able to significantly predict treatment slopes (*mean squared error* (*MSE*) = 19.87, *p* = 9.99e-4, *R*^2^ = 0.21) (Fig. 2A). Describing the correlations between treatment slopes and individual pre-treatment clinical measures, we find that all Vineland and Mullen subscales are significantly positively correlated, while total ADOS score and ADOS RRB was negatively correlated with treatment slopes (Fig. 2B). These results are largely consistent with the idea from past work that pre-treatment clinical measures can be predictive of later treatment outcome (*10, 17*–*22*). However, as a new perspective on this effect, with longitudinal trajectories measured over more than just two time-points (e.g., pre and post-treatment), we find that pre-treatment clinical measures can predict how steep an individual’s treatment slope trajectory will be over the course of the treatment.

**Fig. 2:**
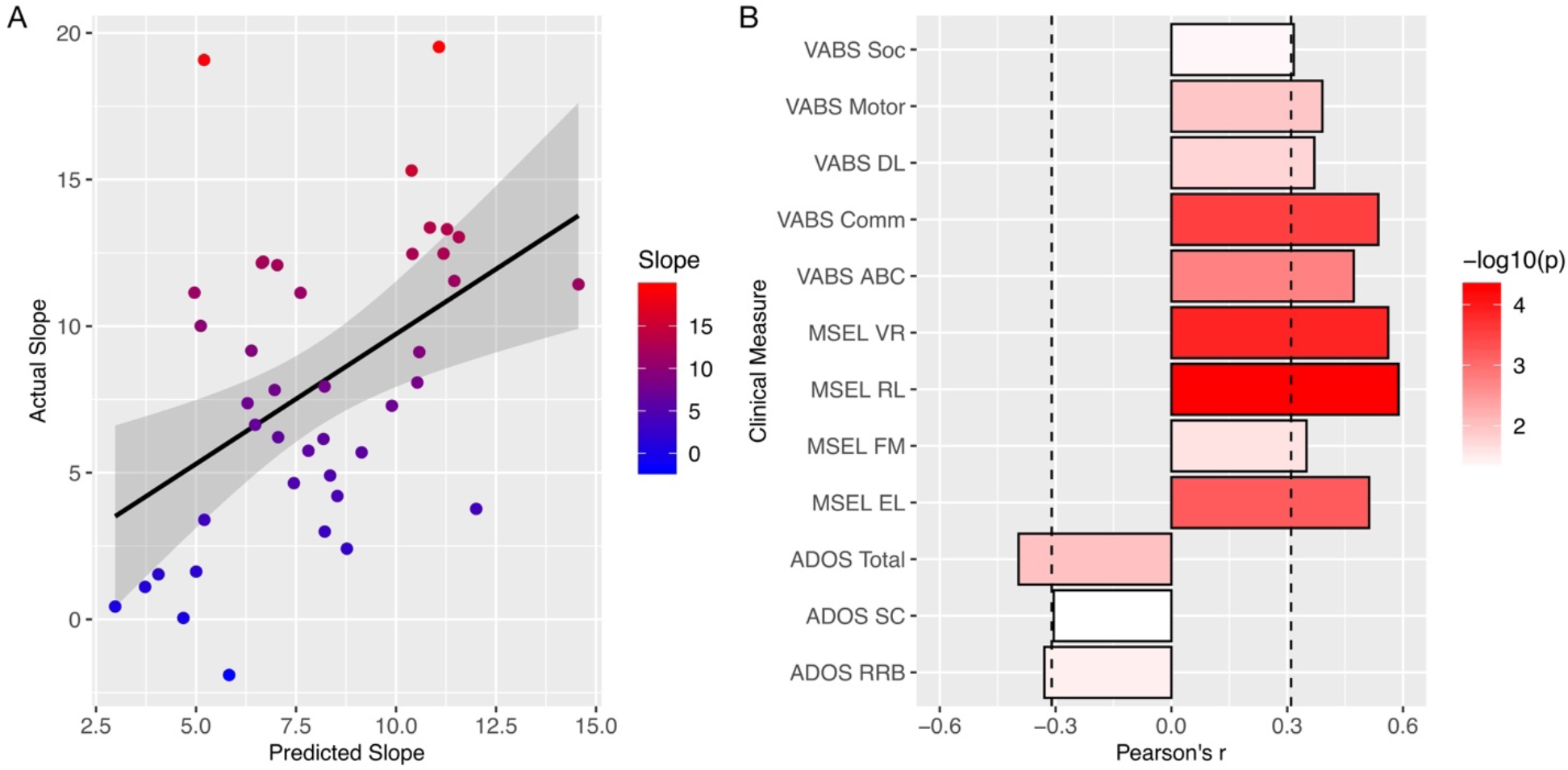
Predicting treatment slopes with pre-treatment clinical measures. Panel A shows actual treatment slopes (y-axis) versus predicted treatment slopes from a LASSO model (x-axis) when using pre-treatment clinical measures as features. Color from blue to red indicates actual treatment slope values. Panel B shows the correlation (Pearson’s r) between treatment slopes and each of the pre-treatment clinical measures. The coloring of the bars indicate the -log10(p-value) and bars that pass the vertical dotted line are measures that pass FDR q<0.05. Abbreviations: ADOS, Autism Diagnostic Observation Schedule; SC, social-communication; RRB, restricted repetitive behaviors; MSEL, Mullen Scales of Early Learning; VR, visual reception; FM, fine motor; RL, receptive language; EL, expressive language; VABS, Vineland Adaptive Behavior Scales; Comm, communication; DL, daily living skills; Soc, socialization; ABC, adaptive behavior composite.

### Prediction of treatment slopes with pre-treatment blood leukocyte gene expression data

We next asked if pre-treatment biological characteristics such as multivariate pre-treatment gene expression in blood leukocytes could also predict treatment slopes. Using a similar LASSO regression approach we find that pre-treatment gene expression can also significantly predict treatment slopes (*MSE* = 21.67, *p* = 0.001, *R*^*2*^ = 0.13) (Fig. 3A), albeit to a lesser extent than pre-treatment clinical behavioral variables (e.g., 13% variance predicted with gene expression versus 21% variance predicted with clinical behavioral measures).

**Fig. 3:**
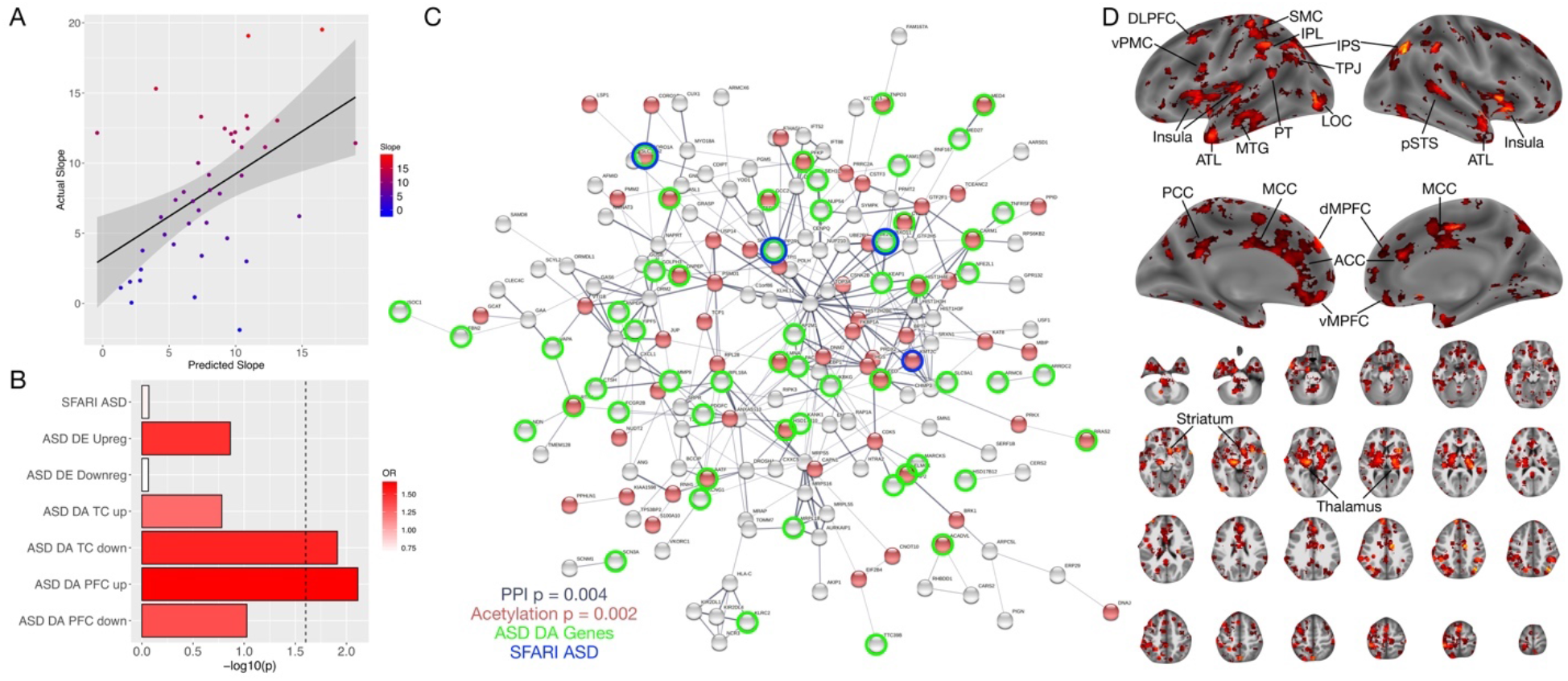
Predicting treatment slopes with pre-treatment blood leukocyte gene expression. Panel A shows actual treatment slopes (y-axis) versus predicted treatment slopes from a LASSO model (x-axis) using pre-treatment blood leukocyte gene expression as features. Color from blue to red indicates actual treatment slope values. Panel B shows the -log10 p-value for the enrichment test (enrichment odds ratio (OR) colored in red) between treatment-relevant genes and ASD-associated gene lists. SFARI ASD refers to genes listed on SFARI Gene (https://gene.sfari.org) where mutations are known to be associated with ASD. DE Upreg or Downreg lists are genes that are differentially expressed (DE) in post-mortem cortical tissue (49). ASD DA lists are genes whose histone proteins are differentially acetylated (DA) in post-mortem cortical tissue (50). Bars passing the dotted line indicate gene lists that pass FDR q<0.05. Panel C shows a graph of the protein-protein interaction (PPI) network of treatment-relevant genes from the LASSO model. Red nodes are genes enriched in UniProt for “acetylation”. Green circles indicate genes whose histone proteins are differentially acetylated (DA) in autism post-mortem cortical tissue. Blue circles indicate genes that high confidence or syndromic ASD genes in SFARI Gene. Panel D shows whole-brain analysis results (thresholded at q<0.05 FDR correction for multiple comparisons) indicating which brain regions show high levels of expression for the treatment-relevant genes. Spatial gene expression was profiled here with the Allen Institute Human Brain atlas. Abbreviations: DE, differentially expressed; DA, differentially acetylated; PFC, prefrontal cortex; TC, temporal cortex; DLPFC, dorsolateral prefrontal cortex; vMPFC, ventromedial prefrontal cortex; dMPFC, dorsomedial prefrontal cortex; ACC, anterior cingulate cortex; MCC, middle cingulate cortex;, PCC, posterior cingulate cortex; vPMC, ventral premotor cortex; PT, planum temporale; TPJ, temporoparietal junction; SMC, somatomotor cortex; IPL, inferior parietal lobule; pSTS, posterior superior temporal sulcus; ATL, anterior temporal lobe; MTG, middle temporal gyrus; LOC, lateral occipital cortex.

Next, we investigated which genes were most important in helping the LASSO model make such treatment slope predictions. Because LASSO uses L1 regularization to shrink coefficients of features that are less informative to 0, this allowed us to identify the subset of key genes that contribute to the model’s predictive accuracy. Here we find that LASSO prioritizes 295 genes that help predict treatment slopes. Rather than being a random array of genes, these treatment-relevant genes show evidence of interactions at the protein level, as evinced with a protein-protein interaction (PPI) analysis (observed edges = 353, expected edges 306, p = 0.004) (Fig. 3C). Further annotation of this treatment-relevant gene set was done with gene set enrichment analysis. This analysis discovered enriched biological processes such as regulation of protein localization and vesicle mediated transport. Cellular compartments such as cytosol, intracellular organelle lumen and cytoplasm were also enriched. With UniProt, we also discovered acetylation as a keyword enrichment (Fig. 3C) (Table 2). Thus, treatment-relevant genes discovered by LASSO likely interact at the protein level and may be involved in processes such as protein localization, vesicle mediated transport, and acetylation.

**Table 2:** Gene Enrichment Analysis. This table shows results of gene set enrichment analysis for treatment-relevant genes. Abbreviations: GO, gene ontology.

| Biological Process (GO) |  |  |  |
| --- | --- | --- | --- |
| GO-term | description | count in network | false discovery rate |
| GO:0032880 | regulation of protein localization | 29 of 901 | 0.0379 |
| GO:0016192 | vesicle-mediated transport | 48 of 1699 | 0.0043 |

| Cellular Component (GO) |  |  |  |
| --- | --- | --- | --- |
| GO-term | description | count in network | false discovery rate |
| GO:0005829 | cytosol | 101 of 4958 | 0.0019 |
| GO:0070013 | intracellular organelle lumen | 98 of 5162 | 0.0190 |
| GO:0005737 | cytoplasm | 186 of 11238 | 0.0037 |

| UniProt Keywords |  |  |  |
| --- | --- | --- | --- |
| keyword | description | count in network | false discovery rate |
| KW-0007 | Acetylation | 74 of 3335 | 0.0024 |

We next asked if this list of treatment-relevant genes might be associated with genetic mutations associated with autism or with genes that show dysregulated expression or histone acetylation in post-mortem cortical tissue. Using gene lists from SFARI Gene (*48*) as well as a list of differentially expressed genes from Gandal et al., (*49*), we find no evidence of enrichment in either of these lists. However, we did find the presence of 4 genes that are either high-confidence and/or syndromic ASD genes in SFARI Gene – *KMT2C, CORO1A, FBXO11*, and *PPP2R5D. KMT2C* is noted as a rare de novo loss of function variant associated with autism (*52*–*57*). *CORO1A* is rare de novo loss of function variant associated with autism (*56*) and is located within the well-known ASD-associated CNV region of 16p11.2 (*58*). *FBXO11* is another rare de novo loss of function and missense variant in autism (*54, 57*) and appears in the autism-associated CNV region of 2p16.3 (*59, 60*). *PPP2R5D* is a known syndromic cause of ASD and rare de novo loss of function variant associated with autism (*56, 61*). Each of these genes are members of the PPI network shown in Fig 3C. Related to the UniProt enrichment in acetylation, we also found significant enrichment with genes that are differentially acetylated in autism post-mortem cortical tissue (Fig. 3B, Table 3). Specifically, treatment-relevant genes were enriched for upregulated histone acetylated genes in prefrontal cortex tissue, but downregulated histone acetylated genes in temporal cortex. This difference in spatial regions and directionality of the histone acetylation effect could suggest that these treatment-relevant genes may asymmetrically impact differing brain regions. Thus, while these treatment-relevant genes map onto a few genes with known evidence for high-confidence mutations or dysregulated gene expression, they are more strongly linked to genes that show evidence of differential histone acetylation in ASD cortical tissue. This potentially indicates that treatment-relevant biology may be linked to epigenetic changes such as histone acetylation. Given that early intervention intends to change behavior through reshaping the underlying biology, these links to acetylation could potentially provide key novel evidence as to how treatment effects may be moderated by individual molecular characteristics intrinsic to each individual.

**Table 3:** Enrichments with ASD-relevant gene lists. This table shows results of enrichment analysis for treatment-relevant genes and ASD-relevant gene lists. Abbreviations: DE, differentially expressed; DA, differentially histone acetylated; OR, enrichment odds ratio; FDR, false discovery rate.

| Gene List | Odds Ratio (OR) | p-value | false discovery rate (FDR) |
| --- | --- | --- | --- |
| SFARI ASD | 0.73 | 0.85 | 0.86 |
| ASD DE Downregulated | 0.68 | 0.86 | 0.86 |
| ASD DE Upregulated | 1.46 | 0.13 | 0.23 |
| ASD DA Prefrontal Cortex Upregulated | 1.64 | 0.007 | 0.04 |
| ASD DA Prefrontal Cortex Downregulated | 1.34 | 0.09 | 0.21 |
| ASD DA Temporal Cortex Upregulated | 1.24 | 0.16 | 0.23 |
| ASD DA Temporal Cortex Downregulated | 1.51 | 0.01 | 0.04 |

Finally, we examined how treatment-relevant genes may be preferentially expressed in different regions of the human brain. Leveraging spatial gene expression information from the Allen Institute Human Brain gene expression atlas, we looked for which regions showed high levels of expression of these treatment-relevant genes. To do this, we downloaded spatial gene expression maps for all 295 treatment-relevant genes from https://neurosynth.org/genes/. With a one-sample t-test in SPM12, we ran a whole-brain analysis to identify brain areas where expression levels were significantly different from 0, correcting for multiple comparisons at voxel-wise FDR q<0.05. Here we find that subcortical areas are highly prominent - particularly the thalamus, striatum, and claustrum. Amongst cortical areas, the most prominent regions are the anterior, middle, and posterior cingulate cortex (ACC, MCC, PC), dorsal and ventral medial prefrontal cortex (dMPFC, vMPFC), dorsolateral prefrontal cortex (DLPFC), ventral premotor cortex (vPMC), somatomotor cortex (SMC), temporoparietal junction (TPJ), planum temporale (PT), inferior parietal lobule (IPL), intraparietal sulcus (IPS), posterior superior temporal sulcus (pSTS), anterior temporal lobe (ATL), middle temporal gyrus (MTG), lateral occipital cortex (LOC), and insular cortex (Ins).

## Discussion

In this work we examined whether pre-treatment clinical behavioral and blood leukocyte gene expression patterns could predict the rate of skill growth in response to early intervention in young toddlers with autism. Congruent with prior studies, pre-treatment clinical behavioral characteristics such as language and communication and non-verbal cognitive ability are indeed helpful for predicting later treatment response (*10, 17*–*22*), predicting around 21% of the variance in treatment slopes. A novel finding from this work is that pre-treatment gene expression patterns from blood leukocytes are also informative for predicting treatment slopes - predicting around 13% of the variance in treatment slopes. The effect of behavioral variables predicting more variance may not be surprising since such variables are conceptually and theoretically closer to what is being measured as the treatment outcome (e.g., behavioral change on the aSLP). However, the effect that pre-treatment blood leukocyte gene expression can predict treatment slopes at all is a revelation, given that prior to this work it was unknown whether pre-treatment biological factors such as blood leukocyte gene expression could predict treatment slopes at all.

Digging deeper into the gene expression signal that is predictive of treatment slopes, our LASSO modeling approach prioritizes a subset of 295 genes that highly interact at the protein level and which are enriched for biological processes such as acetylation. Expanding on the idea of acetylation as a treatment-relevant biological process, we also discovered that these treatment-relevant genes are enriched for genes that are differentially histone acetylated in post-mortem cortical tissue of ASD patients (*50*). Given that the central dogma behind early intervention is to capitalize on an individual’s heightened propensity for neurobiological plasticity and change in early development, these findings suggest that one key to predicting an individual’s propensity for such change may be hidden within individualized and intrinsic biology related to histone acetylation. In other words, predicting early intervention treatment response may hinge critically on how susceptible an individual’s intrinsic biology is to experience- or context-dependent control over regulation of gene expression. This idea bodes well with general ideas regarding histone acetylation as one of the primary molecular influences over activity-dependent gene expression, which would then subsequently alter experience-dependent learning and memory processes (*62*) that are critical ingredients of early intervention.

We also discovered that treatment-relevant genes highly express throughout a range of subcortical and cortical areas. Subcortical areas such as the thalamus, striatum, and claustrum all have extensive connections to the various cortical areas implicated (*63*–*66*). The cortical areas fall within well-known large-scale circuits like the default mode, salience, and somatomotor network. The default mode network is noted for its overlap with regions considered integral for social brain circuitry (e.g., dMPFC, vMPFC, PCC, TPJ, ATL, pSTS) and social-communicative functions linked to the domains affected in ASD (*67*–*75*). Other regions relevant to the mirror system are also apparent (e.g., vPMC, Ins, MCC, IPL, IPS, SMC) (*76*–*78*). Language-relevant regions are also notable, such as (e.g., PT, MTG, vPMC, ATL) (*79, 80*). While speculative, this evidence could be suggestive of the possible impact of treatment-relevant genes on circuitry that plays important roles in cognitive and behavioral domains targeted by early intervention and which are key domains of importance in the early development of autism.

In addition, we also found that starting treatment before versus after 24 months is a meaningful distinction (*10, 11*). Toddlers who started treatment before 24 months showed larger overall gains than those starting treatment after 24 months, even when controlling for the amount of time in treatment. This result is compatible with the main ideas behind why early intervention is crucial before 24 months (*1, 11, 30*). Also compatible with the idea of that treatment start before versus after 24 months is important, we also discovered that treatment slopes are much more variable once past 24 months of age. The enhanced variability of treatment slopes after 24 months is important, as it underscores how heterogeneity can be magnified with later treatment start. One implication of this result is that prediction of treatment outcome is a much more difficult task when the child begins treatment after 24 months of age. This is another consideration for why early detection and intervention is key – treatment outcomes tend to be more consistent if treatment begins before 24 months.

There are some caveats and limitations that are necessary to address to interpret the present findings. First, the results reported here are tied to a specific, evidence-based early intervention program that contains a mixture of elements from various programs (e.g., applied behavioral analysis, pivotal response training) and administered by highly trained providers with systematic probes of fidelity of implementation. The use of the same standardized treatment approach for all participants is a strength of the current study. However, given the variety of different types of early intervention programs available today, caution must be taken in generalizing these findings to other intervention programs. A question for future work would be to examine whether these findings extend to other widely used early intervention programs. Second, the outcome measure operationalized as the rate of improvement over time is not a commonly used metric to evaluate early intervention response. In most designs, there are time-locked pre- and post-testing measures to evaluate treatment response. However, the rate of response to treatment from multiple longitudinal measurements may be a more sensitive measure of treatment response than a change score sampled at just two points in time. Third, these results are identified in a relatively small sample of ASD toddlers. Future work replicating the findings with larger samples is needed. Finally, future work could examine whether different approaches to merge multiple data modalities such as pre-treatment gene expression and clinical measures might help to better predict treatment slopes. In the current work we did not investigate this possibility as it is beyond the scope of the current investigation and requires much more sophisticated approaches tailored specifically for multiple modality data fusion, especially in situations where different modalities are high dimensional and/or differ substantially in dimensionality (*81, 82*).

In conclusion, this work shows the importance of early treatment start ideally before 24 months, and also shows for the first time, that blood gene expression characteristics can predict how fast toddlers with ASD respond to early treatment. While clinical behavioral variables outperformed gene expression measures, the signal within gene expression is important because it potentially indicates that a key biological ingredient for determining an individual’s treatment outcome is susceptibility to epigenetic change via mechanisms such as acetylation. Understanding how this treatment-relevant biology affects neuroplasticity and experience-dependent learning is a key next step towards how such molecular mechanisms are linked to heterogeneous outcomes in ASD.

## Funding and Acknowledgments

We thank all participants and their families for participating in this study. This project has received funding from the European Research Council (ERC) under the European Union’s Horizon 2020 research and innovation programme under grant agreement No 755816 (ERC Starting Grant to MVL). This work was also supported by the following grants to EC, KP, LE, and NEL - NIMH R01-MH080134 (KP), NIMH R01-MH104446 (KP), NFAR grant (KP), NIMH Autism Center of Excellence grant P50-MH081755 (EC, KP), NIMH R01-MH036840 (EC), NIMH R01-MH110558 (EC, NEL), NIMH U01-MH108898 (EC), NIDCD R01-DC016385 (EC, KP, LE, MVL), CDMRP AR130409 (EC), and the Simons Foundation 176540 (EC).

## Author contributions

Conceptualization: MVL, EMB, EC, KP, TP, LS, ACS. Methodology: MVL, TP, EC, KP. Software: MVL, TP, VG. Formal analysis: MVL, EMB, TP, IL, VM, NB. Investigation: LS, ACS, CCB, LL, ECB, EC, KP. Data curation: TP, VG, ECB. Writing - original draft preparation: MVL, EMB. Writing - review and editing: MVL, EMB, EC, KP. Visualization: MVL. Supervision: EC, KP, LS, ACS, MVL. Project administration: EC, KP. Funding acquisition: MVL, EC, KP.

